# Efficacy and Safety of Povidone Iodine versus Bleomycin for Pleurodesis in Malignant Pleural Effusions: A Systematic Review and Meta-Analysis

**DOI:** 10.64898/2026.05.09.26352791

**Authors:** Salamullah, Muhammad, Moh Habib, Sinta Chaira Maulannisa

## Abstract

Malignant pleural effusion (MPE) frequently complicates advanced cancer and impairs quality of life. Chemical pleurodesis with agents such as bleomycin or povidone iodine is widely used, but comparative efficacy and safety remain uncertain. Bleomycin is an established agent but is costly and less available, whereas povidone iodine is affordable and easily accessible. This study aimed to systematically compare the efficacy and safety of bleomycin versus povidone iodine for pleurodesis in patients with malignant pleural effusions.

We conducted a systematic review and meta-analysis following PRISMA guidelines. PubMed, Semantic Scholar, and the Google Scholar were searched through May 20^th^ 2025. Studies included randomized controlled trials and cohort studies comparing bleomycin and povidone iodine for pleurodesis in patients with MPE. Seven studies with 392 patients (174 in the povidone iodine group, 218 in the bleomycin group) were included. Success rates for pleurodesis ranged from 71.1% to 100% for povidone iodine and 66.7% to 95.2% for bleomycin. Meta-analysis showed no significant difference in efficacy (RR = 1.04, 95% CI: 0.94–1.15, p = 0.50; I^2^ = 43%). Both agents were well tolerated, with similar rates of mild adverse events. This study showed no significant bias.

Povidone iodine and bleomycin are equally effective and safe for pleurodesis in MPE. Given its lower cost and greater accessibility, povidone iodine may be preferred, especially in resource-limited settings.

## INTRODUCTION

Malignant pleural effusion (MPE) is a common and debilitating complication in patients with advanced malignancies, particularly lung and breast cancers. It is estimated that MPE affects up to 15% of all patients with cancer, leading to significant morbidity, reduced quality of life, and limited survival [1]. The primary goals of management are to alleviate symptoms such as dyspnea and prevent the recurrence of fluid accumulation.

Chemical pleurodesis is widely used as a palliative intervention for recurrent MPE, promoting adhesion between the parietal and visceral pleura to obliterate the pleural space [2]. Multiple sclerosing agents have been investigated for this purpose, including talc, tetracyclines, bleomycin, and povidone iodine [3,4]. Bleomycin, an antineoplastic antibiotic, is among the established agents for pleurodesis and is included in many guidelines. However, its use is often limited by high cost, restricted availability in some regions, and potential systemic toxicity [5].

Povidone iodine is a widely available, inexpensive antiseptic with recognized efficacy as a sclerosing agent for pleurodesis [6]. It has been increasingly adopted in resource-limited settings due to its accessibility and low cost. Despite multiple individual studies comparing povidone iodine to bleomycin for pleurodesis, there remains uncertainty regarding their relative efficacy and safety.

To address this gap, we performed a systematic review and meta-analysis to compare the efficacy and safety of bleomycin versus povidone iodine for pleurodesis in patients with MPEs.

## METHODS

### Search Strategy and Study Selection

This systematic review and meta-analysis were conducted according to the preferred reporting items for systematic reviews and meta-analyses (PRISMA) guidelines. A comprehensive literature search was performed in PubMed, Semantic Scholar, and the Google Scholar, using keywords terms: “pleurodesis,” “malignant pleural effusion,” “bleomycin,” and “povidone iodine.” Additional studies were identified by reviewing references of included articles and relevant reviews according to our PICO (population, intervention, comparison, outcome) design (table 1).

**Table 1.** PICO design of study.

| Component | Description |
| --- | --- |
| Population | Patients with malignant pleural effusions |
| Intervention | Pleurodesis using povidone iodine |
| Comparison | Pleurodesis using bleomycin |
| Outcome | Primary: Success of pleurodesis<br>Secondary: Adverse events |

**Table 2.** Summary of included studies comparing povidone iodine and bleomycin for pleurodesis in patients with MPE. The table lists study design, country, patient demographics, cancer types, success rates, outcome definitions, follow-up periods, adverse events, and key conclusions. Success rates are presented as the number of patients achieving successful pleurodesis over the total number treated (n/N) and as percentages for each agent. PI = povidone iodine; B = bleomycin

| No. | Study | Design, Country | Age (Mean/Range) | Bleomycin n/N, Success % | Povidone Iodine n/N, Success % | Outcome Definition | Follow-up | Adverse Events | Notes/Conclusions |
| --- | --- | --- | --- | --- | --- | --- | --- | --- | --- |
| 1 | **Aboalazayem 2023** | Prospective, Egypt | 45–76 (mean 51±7) | 51/60 (85.0%) | 32/45 (71.1%) | Drain <100 ml/day + X-ray | Not stated | Not stated | Both Bleo and PI effective |
| 2 | **Asghar 2011** | RCT, Iran | Not stated | 15/19 (79%) | 15/20 (75%) | Complete response (symptom+X-ray) | 1 month | Lower dyspnea in bleo group | Similar efficacy, PI cheaper |
| 3 | **Bagheri 2018** | RCT, Iran | B: 58.0±9.3, PI: 59.6±7.7 | 20/30 (66.7%) | 25/30 (83.3%) | No recurrence 6 mo (X-ray) | 6 months | Chest pain, fever, hypotension, equal in both groups | PI as effective & cheap |
| 4 | **Bakr 2012** | Prospective, Egypt | B: 45–73 (60.7), PI: 46–71 (64.3) | 7/10 (70%) | 8/10 (80%) | No reaccumulation 1 wk & 3 mo | 3 months | Not stated | PI as effective & cheap |
| 5 | **Fahad 2021** | Retrospective, Iraq | B: 58±10.9, PI: 62±11.3 | 26/33 (78.8) | 20/21 (95.2%) | Recurrence at 3 months | 3 months | Chest pain (B:2, PI:3), Fever (B:2) | Recurrence: B: 21.2%, PI: 4.8% |
| 6 | **Murni 2023** | Prospective, Indonesia | Not stated | 19/23 (82.6%) | 23/23 (100%) | WSD <100 ml/24h + lung expansion (X-ray) | Not stated | VAS pain; not detailed | No significant difference |
| 7 | **Saleh 2020** | Retrospective, Egypt | 22–74 (mean 57.5) | 34/43 (79.1%) | 19/25 (76%) | Prevent reaccumulation, symptoms | ≥1 month | Not stated | All agents safe, efficacy driven by access/cost |

Studies were eligible if they:

- Included patients with MPEs;
- Compared bleomycin versus povidone iodine as agents for chemical pleurodesis;
- Reported on at least one of the following outcomes: pleurodesis success, recurrence, or adverse events;
- Were randomized controlled trials (RCTs), prospective, or retrospective cohort studies.

Two reviewers independently screened titles and abstracts, assessed full-text articles for eligibility, and extracted data. Disagreements were resolved by discussion.

### Data Extraction and Quality Assessment

The following data were extracted from each study: author, year, country, study design, number of participants, patient characteristics, type of cancer, intervention details, pleurodesis success rates, definitions of outcomes, follow-up duration, and reported adverse events. Study quality and risk of bias were assessed using the funnel plot.

### Outcomes

The primary outcome was the success of pleurodesis, defined as the absence of recurrent effusion based on clinical assessment and/or imaging, at the last follow-up. Secondary outcomes included recurrence rates, adverse events (e.g., chest pain, fever), and mortality where reported.

### Statistical Analysis

Meta-analysis was performed using the fixed method to pool risk ratios (RRs) for pleurodesis success and adverse events, with 95% confidence intervals (CIs). Heterogeneity was assessed using the Chi^2^ test and quantified by the I^2^ statistic. A random-effects model was applied due to anticipated heterogeneity in study populations and protocols. Statistical significance was set at p < 0.05. All analyses were conducted using Review Manager (RevMan) version 5.3.

## RESULTS

### Study selection and characteristics

The PRISMA flow diagram (figure 1) illustrates the study selection process. Of 710 records identified, 521 duplicates were removed, leaving 197 records for screening. After title and abstract screening, 34 articles were assessed for eligibility. Eight full-text articles matched the PICO criteria, but one was excluded due to lack of full-text availability. Finally, seven full-text articles were included in the quantitative synthesis (meta-analysis).

**Fig 1.**
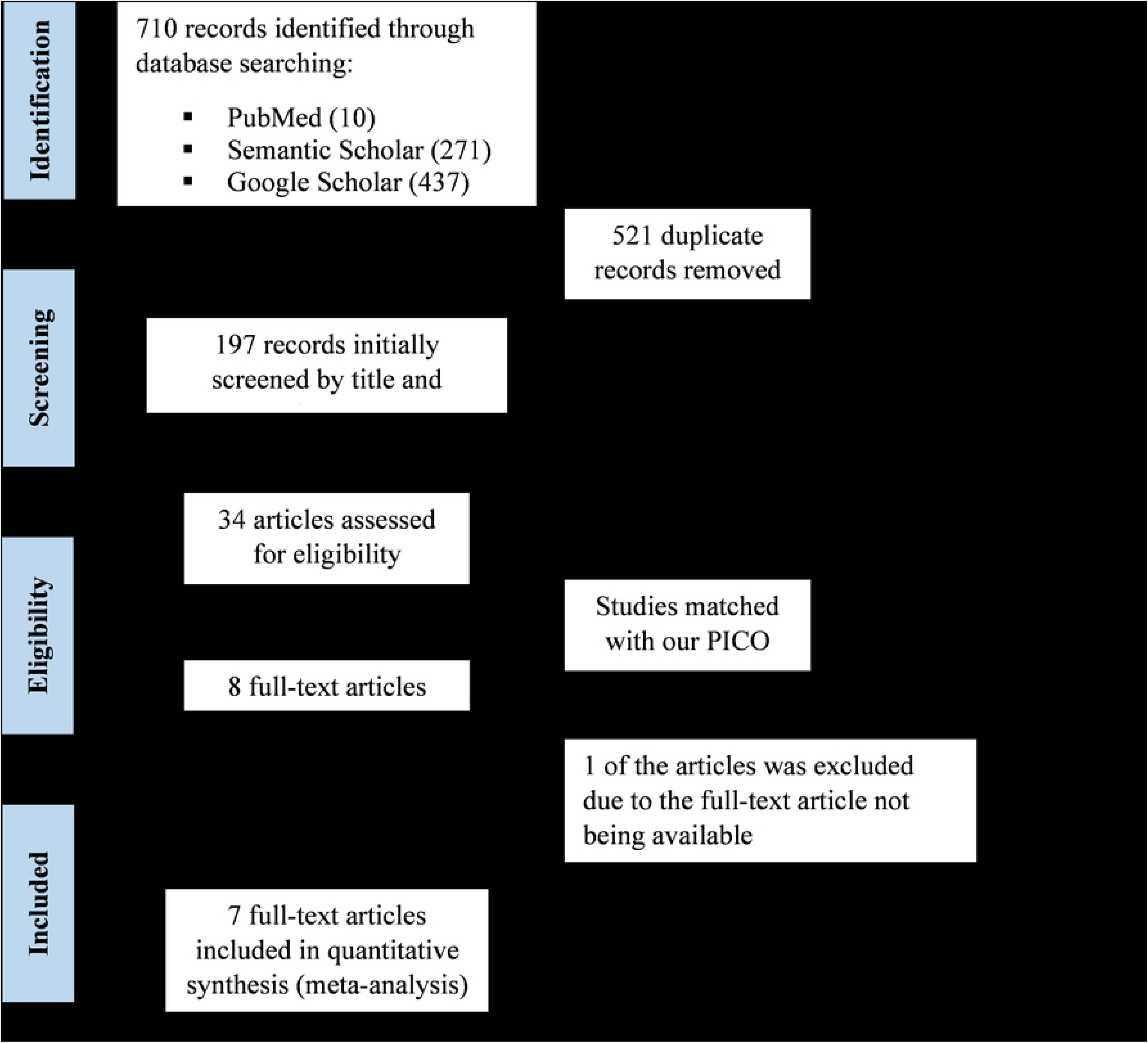
PRISMA flow diagram showing the selection process for studies included in the meta-analysis.

A total of seven studies, comprising 392 patients (174 in the povidone iodine group and 218 in the bleomycin group), were included in the meta-analysis. Study designs included randomized controlled trials and prospective or retrospective cohort studies, conducted across Iran, Egypt, Iraq, and Indonesia [1-4],[7-9]. The baseline characteristics of included studies are summarized in table 1.

### Efficacy of pleurodesis

The individual study success rates for pleurodesis ranged from 71.1% to 100% for povidone iodine and 66.7% to 95.2% for bleomycin. The pooled analysis demonstrated no statistically significant difference in pleurodesis success between povidone iodine and bleomycin (Risk Ratio [RR] = 1.04, 95% CI: 0.94–1.15, p = 0.50; figure 2).

**Fig 2.**
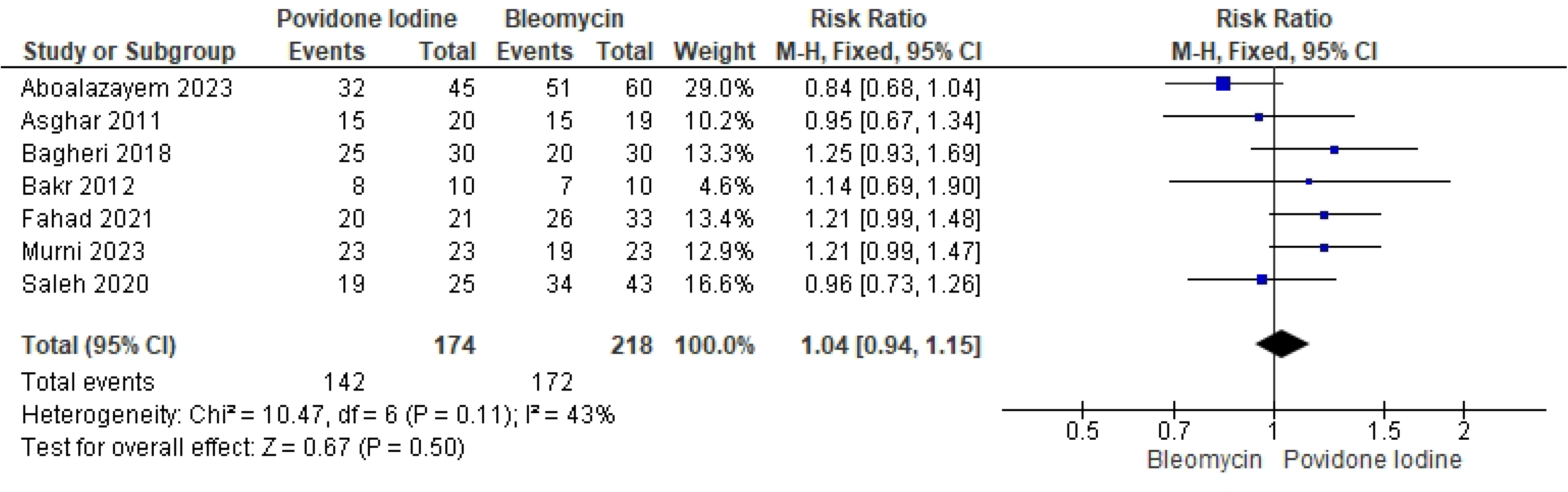
Forest plot of the pooled risk ratio (RR) and 95% confidence intervals (CI) comparing the efficacy of povidone iodine and bleomycin for pleurodesis in patients with MPE. Squares represent the effect size for each included study, with the size proportional to the study’s weight. Horizontal lines indicate 95% CIs. The diamond represents the overall pooled estimate. A risk ratio greater than 1 favors povidone iodine, while a risk ratio less than 1 favors bleomycin. The vertical line indicates no effect (RR = 1).

### Heterogeneity

Moderate heterogeneity was observed among the studies (I^2^ = 43%). The test for overall effect was not significant (Z = 0.67, p = 0.50).

### Adverse events

Analysis of reported adverse events showed that both agents were generally well tolerated. The most common complications were mild and included transient chest pain and fever. No significant differences in adverse event rates were identified between povidone iodine and bleomycin across the included studies.

### Risk of bias assessment

Funnel plot analysis (figure 3) showed an approximately symmetrical distribution of studies on both sides of the pooled effect estimate, indicating no significant publication bias in this meta-analysis. Most studies were evenly spread within the expected funnel-shaped area, and there were no major gaps or clustering.

**Fig 3.**
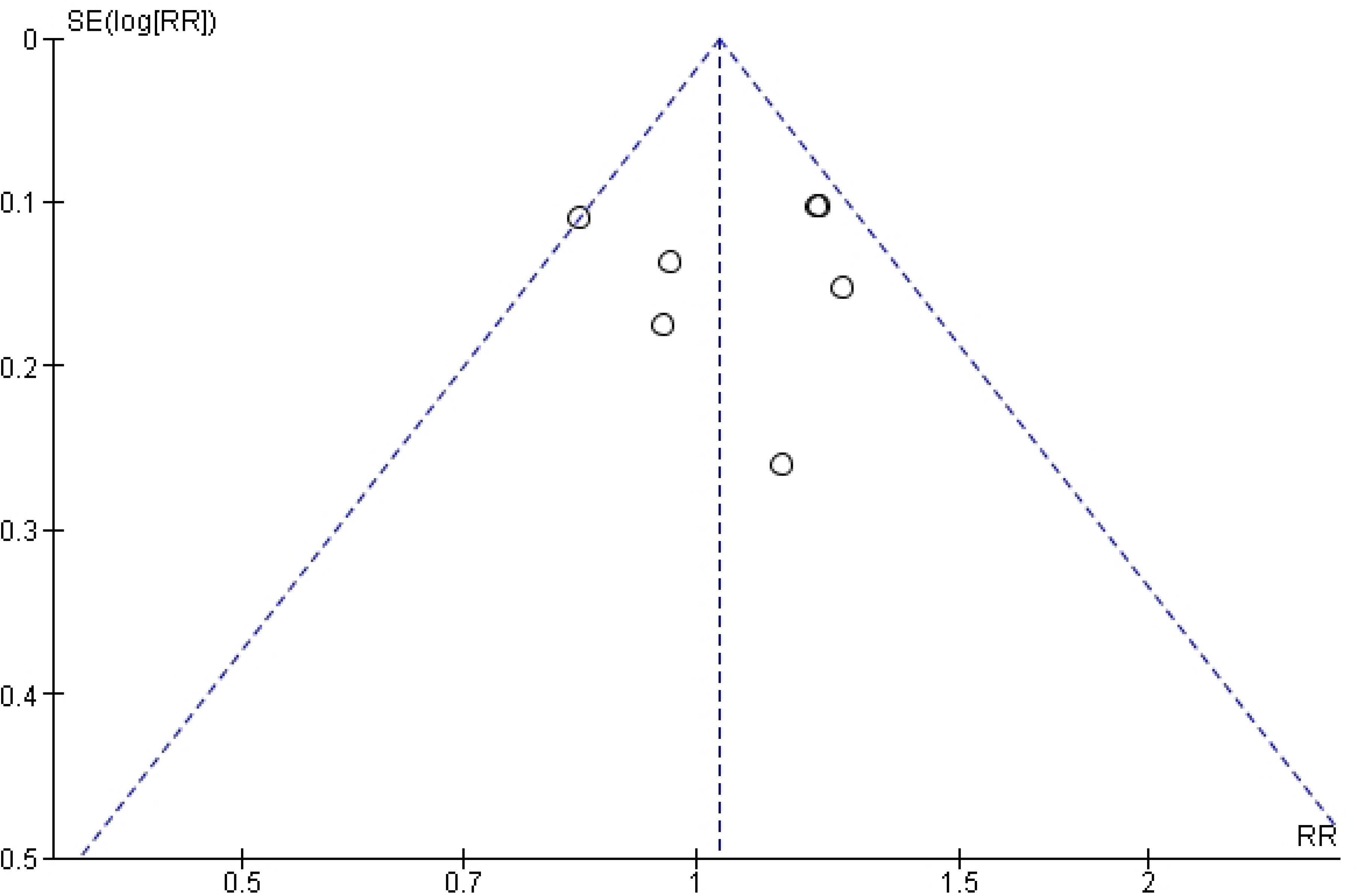
Funnel plot for assessment of publication bias. Each dot represents an individual study; the vertical line indicates the pooled effect estimate. The symmetric distribution suggests no significant publication bias.

Although the number of included studies is relatively small, which can limit the ability to detect subtle asymmetry, the visual findings suggest that the pooled results are robust. Overall, there is no strong evidence that publication or small-study bias influenced the conclusions of this analysis.

## DISCUSSION

This meta-analysis demonstrates that povidone iodine is as effective and safe as bleomycin for pleurodesis in patients with malignant or recurrent MPE. The pooled risk ratio did not show a statistically significant difference, and success rates were similar across all included studies. Both agents achieved high rates of successful pleurodesis, and the risk of adverse events was low and comparable between groups. This results are consistent across various studies and settings, indicating the robustness of the findings.

These findings are supported by previous individual studies. For example, Bagheri et al. found no statistically significant difference in pleurodesis success rates between povidone iodine (83.3%) and bleomycin (66.7%) in their randomized controlled trial [1]. Similarly, Asghar et al. reported similar efficacy for both agents in an RCT, with success rates of 75% for povidone iodine and 79% for bleomycin [2]. Murni et al. also observed no significant difference, with both agents demonstrating high success rates in a cohort from Indonesia [3]. Other recent studies, such as Fahad et al., reported even higher success rates for povidone iodine (95.2%) compared to bleomycin (78.8%), highlighting the potential advantages of povidone iodine in certain patient populations [4].

Several previous systematic reviews and meta-analyses have also highlighted the efficacy and safety profile of povidone iodine as a sclerosing agent. International guidelines and reviews have also recognized povidone iodine as a practical and cost-effective agent for pleurodesis, especially in low- and middle-income countries where access to bleomycin may be limited [5,6]. Agarwal et al. (2012) included 13 studies with 499 patients and found a pooled success rate of 88.7% (95% CI: 84.1–92.1) for iodopovidone pleurodesis, with chest pain being the most common complication and no reported deaths [10]. Similar findings were observed in another systematic review by Agarwal et al. (2006), which reported an overall success rate of 90.6% for iodopovidone, independent of the procedure used or the indication for pleurodesi [11].

Additional single-center studies further support the effectiveness of povidone iodine. Kahrom et al. found an efficacy of 82.2% with povidone-iodine pleurodesis in 63 MPE patients, with only minor side effects and no significant changes in thyroid or renal function [12]. Godazandeh et al. also reported a high overall success rate of 91.6% for povidone-iodine, with pain being the most frequent adverse event and no significant impact on thyroid function [13].

In comparative studies, povidone iodine has shown similar efficacy to other widely used agents such as talc and bleomycin. Mohsen et al. conducted a randomized controlled trial comparing povidone-iodine with thoracoscopic talc poudrage in breast cancer patients and found no significant difference in recurrence rates or overall efficacy, but noted a shorter hospital stay with povidone iodine [14]. Ibrahim et al. found no significant difference in recurrence or complication rates between povidone-iodine and talc pleurodesis, supporting its use as a cost-effective and safe alternative, especially in resource-limited settings [15].

When compared with bleomycin, multiple studies have demonstrated equivalence in efficacy and safety. Shaw and Agarwal’s Cochrane review established that chemical sclerosants, including bleomycin and povidone iodine, are both effective for pleurodesis, though talc appears to be the most effective agent overal [16] However, in many countries, talc is either unavailable or its use is restricted due to concerns over complications such as acute respiratory distress syndrome (ARDS), making povidone iodine and bleomycin practical alternative [17].

It is important to note that while bleomycin is recommended in some guidelines and is widely used, its higher cost and limited availability are significant barriers, particularly in low- and middle-income countries. In contrast, povidone iodine is inexpensive, accessible, and has demonstrated similar clinical effectiveness, making it a viable first-line agent for pleurodesis worldwide.

While the overall evidence supports the use of povidone iodine, there are still areas requiring further investigation. Variations in patient selection, technique, and definitions of success across studies introduce some heterogeneity. Most adverse events associated with povidone iodine are mild and self-limiting, with chest pain and fever being most commonly reported. Importantly, no procedure-related mortality has been consistently reported in the literature.

In summary, the cumulative evidence confirms that povidone iodine is as effective and safe as bleomycin for pleurodesis in MPE. Given its low cost, broad availability, and favorable safety profile, povidone iodine should be considered an excellent alternative to bleomycin, especially in settings where access to more expensive agents is limited.

Moderate heterogeneity (I^2^ = 43%) was observed in the present analysis. This may be attributed to variations in study design, patient populations, underlying malignancy types, dosing regimens, procedural techniques, and definitions of pleurodesis success across studies [1-7].

Despite these differences, the overall consistency in direction and magnitude of effect across diverse settings strengthens the robustness of our findings.

Limitations of this meta-analysis include the relatively small sample sizes of some studies, limited reporting of long-term outcomes and adverse events, and variability in study design. Most studies were single-center and not all were randomized controlled trials, which may introduce bias. Nevertheless, the pooled evidence consistently indicates equivalence in efficacy and safety between povidone iodine and bleomycin for pleurodesis in MPE.

## CONCLUSION

Povidone iodine and bleomycin are equally effective and safe for pleurodesis in MPE. Povidone iodine is cheaper and more accessible, making it a practical alternative, especially in resource-limited settings. Further high-quality randomized controlled trials are encouraged to confirm these findings and to optimize pleurodesis protocols.

## Data Availability

All relevant data are within the manuscript and its Supporting Information files. The study is based on previously published data, all of which are cited within the manuscript.

## DECLARATIONS

### Ethical approval

Ethical approval was not required for this study as it was a systematic review and meta-analysis.

### Human and animal rights

This article does not contain any studies with animals performed by any of the authors.

### Informed consent

For this type of study formal consent is not required.

### Author contributions

**Conceptualization:** Salamullah, Muhammad

**Data curation:** Salamullah, Muhammad

**Formal analysis:** Moh Habib, Salamullah, Muhammad

**Methodology:** Moh Habib

**Validation:** Sinta Chaira Maulannisa

**Visualization:** Moh Habib, Sinta Chaira Maulannisa

**Writing – original draft:** Sinta Chaira Maulannisa

**Writing – review & editing:** Salamullah, Muhammad, Moh Habib, Sinta Chaira Maulannisa

## REFERENCES

1. Bagheri R, Noori M, Rajayi M, Attaran D, Mohammad Hashem Asna Ashari A, Mohammadzadeh Lari S, Basiri R, Rezaeetalab F, Afghani R, Salehi M. The effect of iodopovidone versus bleomycin in chemical pleurodesis. Asian Cardiovasc Thorac Ann. 2018 Jun;26(5):382–386. 10.1177/0218492318778485.

2. Alavi AA, Eshraghi M, Rahim MB, Meysami AP, Morteza A, Hajian H. Povidone-iodine and bleomycin in the management of malignant pleural effusion. Acta Med Iran. 2011;49(9):584–587. PMID: 22052141

3. Murni TW, Maryani E, Rahantan TS. Comparison of the Effectiveness between Povidone-Iodine and Bleomycin as Pleurodesis Agents in Patients with Malignant Pleural Effusion at Dr. Hasan Sadikin General Hospital, Bandung, Indonesia. Bioscientia Medicina. 2023;7(3):3221–3228. 10.37275/bsm.v7i3.802

4. Macklin PS, et al. The future of chemical pleurodesis: A review of novel and investigational sclerosant agents. Am J Med Sci. 2024;368(3):175–175. 10.1016/j.amjms.2024.04.008

5. Agarwal R, Aggarwal AN, Gupta D. Efficacy and safety of povidone iodine for chemical pleurodesis: a systematic review and meta-analysis. Respirology. 2010;15(4):594–599. 10.1111/j.1440-1843.2009.01663.x

6. Roberts ME, Neville E, Berrisford RG, Antunes G, Ali NJ; British Thoracic Society Pleural Disease Guideline Group. Management of a malignant pleural effusion: British Thoracic Society Pleural Disease Guideline 2010. Thorax. 2010;65(Suppl 2):ii32–ii40. 10.1136/thx.2010.136994

7. Aboalazayem HM, Elfeky MS. Comparative study between different agents used in chemical pleurodesis. Egypt J Hosp Med. 2024 Jan;94(1):1000–2. 10.21608/ejhm.2024.345509

8. Saleh ME, Awad G, Sanad M. Chemical pleurodesis for malignant pleural effusion: which agent is perfect? Cardiothorac Surg. 2020;28(1):12. doi:10.1186/s43057-020-00022-3. https://doi.org/10.1186/s43057-020-00022-3

9. Bakr RM, El-Mahalawy II, Abdel-Aal GA, Mabrouk AA, Ali AA. Pleurodesis using different agents in malignant pleural effusion. Egypt J Chest Dis Tuberc. 2012;61(4):399–404. http://dx.doi.org/10.1016/j.ejcdt.2012.07.005

10. Agarwal R, Singh N, Gupta D. Efficacy & safety of iodopovidone pleurodesis: a systematic review & meta-analysis. Indian J Med Res. 2012;135:297–304. PMID: 22561614

11. Agarwal R, Aggarwal AN, Gupta D. Efficacy and safety of iodopovidone pleurodesis: a meta-analysis. Respirology. 2006;11(5):589–594. 10.1016/j.rmed.2006.02.009

12. Kahrom M, Kahrom H, Mirmohammad Sadeghi A. Efficacy and Safety of Iodopovidone Pleurodesis. Indian J Palliat Care. 2017;23(1):53–57. 10.4103/0973-1075.197958

13. Godazandeh G, Qasemi NH, Saghafi M, Mortazian M, Tayebi P. Pleurodesis with povidone-iodine, as an effective procedure in management of patients with malignant pleural effusion. J Thorac Dis. 2013;5(2):141–141. doi:10.3978/j.issn.2072-1439.2013.02.02. https://doi.org/10.3978/j.issn.2072-1439.2013.02.02

14. Mohsen TA, Zeid AA, Meshref M, Tawfeek S, Ghalli A, Amer K, et al. A prospective randomized trial comparing povidone-iodine and talc pleurodesis in breast carcinoma patients. Eur J Cardiothorac Surg. 2011;40(2):282–286. 10.1016/j.ejcts.2010.09.005

15. Ibrahim IM, Dokhan AL, El-Sessy AA, Eltaweel MF. Povidone-iodine pleurodesis versus talc pleurodesis in preventing recurrence of malignant pleural effusion. J Cardiothorac Surg. 2015;10:64. 10.1186/s13019-015-0270-5

16. Shaw P, Agarwal R. Pleurodesis for malignant pleural effusions. Cochrane Database Syst Rev. 2004;(1):CD002916. 10.1002/14651858.cd002916.pub2

17. Astoul P. Pleurodesis for recurrent malignant pleural effusions: the quest for the Holy Grail. Eur J Cardiothorac Surg. 2011;40(2):277–279. 10.1016/j.ejcts.2010.11.035

